# Metabolic factors associated with plasma phosphorylated tau analytes used to detect Alzheimer’s disease pathology

**DOI:** 10.1101/2025.07.27.25332280

**Authors:** Kao Lee Yang, Yue Ma, Gilda Ennis, Erin Jonaitis, Ramiro Eduardo Rea Reyes, Sterling C. Johnson, Nicholas J. Ashton, Kaj Blennow, Henrik Zetterberg, Barbara B Bendlin

**Author notes:** **Corresponding Author:** Barbara Bendlin, PhD; 600 Highland Ave J5/1 Mezzanine, Madison, WI 53792 USA.

## Abstract

**INTRODUCTION:** The interpretation of plasma phosphorylated tau (pTau) levels may be influenced by metabolic conditions, such as insulin resistance (IR), type 2 diabetes mellitus (T2DM), obesity, and kidney function. We examined the extent to which metabolic factors are associated with plasma pTau concentrations (pTau181, pTau217, pTau231) and the contribution of these factors on analytical outcomes.

**METHODS:** We analyzed data from 287 participants using partial Spearman’s rho, Mann Whitney U, ROC analysis, and linear mixed models.

**RESULTS:** Concordance between plasma pTau217 and amyloid PET was not influenced by IR or estimated-glomerular-filtration-rate (eGFR). Accounting for eGFR improved concordance of pTau181 and pTau231 with amyloid PET. Age, binarized waist-to-hip ratio, and amyloid status were associated with all longitudinal pTau concentrations, and eGFR associated with longitudinal pTau231.

**DISCUSSION:** Metabolic health may influence the interpretation of some plasma pTau biomarkers, potentially necessitating adjustments for metabolic factors in research and clinical settings.

## 1. INTRODUCTION

Alzheimer’s disease (AD) is characterized by an accumulation of amyloid plaques and neurofibrillary tangles formed of hyperphosphorylated tau protein in the brain^1^. These features can be measured with imaging and fluid biomarkers, including measures of secreted phosphorylated tau (pTau) in plasma^2^. Plasma measurements of pTau at amino acid residues 181, 217, and 231 are strongly concordant with amyloid positron emission tomography (PET) assessments^3–6^, with pTau217 emerging as the most promising plasma biomarker for AD pathology. Although pTau analytes can be used to support AD dementia diagnosis, the extent to which chronic metabolic health conditions may impact pTau measurements is not fully known. This is important given that conditions such insulin resistance (IR), type 2 diabetes mellitus (T2DM), obesity, and kidney function may impact plasma levels by altering tau phosphorylation^7–9^, disrupting blood-brain barrier integrity^10^, or impairing protein clearance^11,12^.

Here we sought to determine the extent to which metabolic factors, including IR, T2DM, obesity, and kidney function are associated with plasma pTau (181, 217, and 231) biomarker concentrations. For the conditions found to be associated with pTau, we further examined whether accounting for these factors could impact analytical outcomes. For a subset of individuals with serial plasma measurements, we investigated the relationships of these metabolic conditions with the mean level of pTau analytes. We hypothesized that IR, T2DM, obesity, and kidney function would be associated with plasma pTau analytes and that accounting for these would improve the concordance between plasma pTau and brain amyloid burden as measured with PET.

## 2. METHODS

### 2.1 Participants

This analysis drew from a subsample of participants enrolled in the Wisconsin Registry for Alzheimer’s Prevention (WRAP) study^13^ with available serial plasma data for all pTau analytes: pTau181, pTau217, and pTau231 (n=425). Of these participants, 5 were excluded for missing insulin and glucose data (to calculate IR values), and 133 were excluded for having an amyloid PET scan that was acquired greater than 2.5 years from the plasma acquisition for a final sample size of 287 participants. All study procedures were approved by the University of Wisconsin-Madison’s Institutional Review Board.

### 2.2 Plasma and amyloid PET acquisition

All participants underwent a blood draw following 8-12 hour fast. Plasma pTau181 and pTau231 concentrations were measured using Single molecule array (Simoa) assays developed at the University of Gothenburg on HD-X instruments (Quanterix, Billerica, MA)^3,4^. Plasma pTau217 was measured using the ALZPath pTau217 Simoa assay on Quanterix HD-X as previously described^6^. Brain amyloid was quantified through [C-11]PiB PET scans. Pet scans were processed using an inhouse pipeline and global PiB DVR (Logan graphical analysis, cerebellar gray matter reference region) was derived by averaging across 8 bilateral regions of interest. Participants were dichotomized into beta-amyloid (Αβ) positive versus negative groups based on a previously derived in-house cutoff of PiB DVR>1.19^14^.

### 2.3 Metabolic measurements

#### 2.3.1 Insulin Resistance

IR was measured using the updated homeostatic model assessment of IR (HOMA2-IR) and z-scored. HOMA2-IR^15^ is an improved version of the original HOMA-IR^16^ metric because it accounts for peripheral insulin secretion variations that may be influenced by hepatic function^17^.

#### 2.3.2 Diabetes Status

Diabetes status (positive or negative) at all visits were determined using fasting blood glucose ≥ 126 (mg/dL) and/or medication usage to treat T2DM (e.g., Metformin). The HOMA model was derived from normoglycemic and mildly diet-treated (not medication treated) individuals with T2DM^16^. Given that T2DM was defined with medication usage, IR was not calculated for individuals with T2DM at initial visit or at follow-up visits where individuals reported developing T2DM to avoid potential confounding.

#### 2.3.3 Obesity

BMI has been commonly used as a metric of obesity^18^. However, the Lancet Diabetes & Endocrinology Commission on Clinical Obesity published guidelines in 2025 stating that BMI use be restricted to population screening purposes and advised that central measures, such as waist-to-hip ratio (WHR), be used to accurately assess excess adiposity (Rubino et al^19^). As such, for this study, we calculated both general (BMI) and central (WHR) obesity measurements. We generated binarized classifications for BMI and WHR based on established cutoffs. BMI was classified as obese (≥30 kg/m²) or non-obese (<30 kg/m²), and WHR was classified as high (men: >0.90, women: >0.85) or normal (men: ≤0.90, women: ≤0.85)^19^.

#### 2.3.4 Kidney Function

Kidney function was assessed using the estimated glomerular filtration rate (eGFR), derived from serum creatinine levels (mg/dL), according to the 2021 Chronic Kidney Disease – Epidemiology Collaboration (CKD-EPI) equation^20^. We computed the eGFR for the subset of individuals in this study that had creatinine values at the time of plasma data collection (n=73). For the subset with eGFR data, 37 individuals met the criteria for mildly decreased kidney function (eGFR 60-89 mL/min/1.73 m^2^). However, since no individual met the criteria for chronic kidney disease (eGFR < 60 mL/min/1.73 m^2^)^20^, we utilized eGFR analytically to indicate normal to mildly decreased kidney function on a continuous scale with lower eGFR values indicating worse kidney function and higher values indicating better kidney function^20^.

### 2.4 Measurement Schedule

Plasma pTau analytes were the primary outcome measures with 142 individuals having a single measurement, while others had up to four measurements available. For those with longitudinal data, plasma measurements were acquired 3.43 ± 0.77 years apart (mean ± SD), with the shortest time interval being 1.78 years and the longest interval being 6.78 years. Given that not every available plasma pTau datapoint had a corresponding PET scan and/or available glucose and insulin data, this spacing in years between plasma data measurements does not reflect the fixed 2-year WRAP schedule^13^. All HOMA2-IR, WHR, BMI, assessments for T2DM, and creatinine measurements were acquired at the time of plasma measurement, except for amyloid PET where we utilized the scan acquired closest to the plasma measurement (mean ± SD time interval: 0.81 ± 0.77 years).

### 2.5 Statistical Analysis

#### 2.5.1 Cross-sectional analysis

Cross-sectional analyses were performed using data from the time of the first plasma measurement. To evaluate the relationship between pTau analyte levels among binarized variables (diabetes status, BMI, and WHR), we first assessed normality of data distribution and homogeneity of variances. Shapiro-Wilks test showed that all three pTau analytes (pTau181, pTau217, and pTau231) significantly deviated from normality, indicating violation of the normality assumption for parametric tests. Next, Levene’s test showed that all pTau analytes met the assumption for equal variances, except for pTau217 among the BMI groups. Given these results, we compared pTau analytes across binarized groups using the non-parametric Mann-Whitney U test to assess median differences in pTau concentration between groups. For continuous variables HOMA2-IR and eGFR, we used partial Spearman’s rho correlations to test the strength and direction of relationships with pTau analytes at baseline measurements adjusting for age. Any metabolic factor that indicated a weak or non-existent relationship (p>.05) in the prior described analyses were excluded from receiver operating characteristic (ROC) analysis.

We next performed ROC analysis to evaluate how well each plasma pTau biomarker can correctly classify brain amyloid PET positivity when metabolic factors are accounted for. To perform ROC analysis, we first conducted logistic regression models predicting Αβ positive or negative (Αβ+/-) PET status not accounting for metabolic factors (IR or eGFR) significantly related to pTau analytes (Model1), accounting for IR (Model 2), or eGFR (Model 3). Model 1 included pTau (181, 217, or 231), age at plasma acquisition, and gender as predictors to classify Αβ+/- PET. Model 2 included all Model 1 predictors and IR. Model 3 included all Model 1 predictors and eGFR. Predicted probabilities of being amyloid PET positive were obtained from these models. These estimates were then used as a continuous predictor to derive area under the ROC curve (AUC) and 95% confidence intervals to assess model discrimination.

#### 2.5.2 Longitudinal analysis

To examine the longitudinal association between metabolic factors and plasma pTau measurements, we fitted three linear mixed-effects models - one for each of the pTau analytes (pTau181, pTau217, and pTau231) – that included random intercepts and fixed effects for age (mean-centered), gender, amyloid positivity status, z-scored HOMA2-IR, and binarized WHR. As a sensitivity analysis, we also ran all primary models using z-scored WHR as a continuous variable. We did not include T2DM in these models as we were underpowered to reliably detect an effect of T2DM given the small number of individuals who had the disease. We selected WHR over BMI as the primary obesity measure as prior studies have shown this metric to have greater relevance to dementia risk and cognitive impairment^21–23^. Given the limited number of people with eGFR data, we first performed model selection on a subset with complete data (N=70) using AIC and BIC to select the best model for each pTau analyte. For pTau181, models without eGFR (AIC = 212.93, BIC = 231.89) performed better than models with eGFR (AIC = 214.19, BIC = 235.51). Similarly, pTau217 models without eGFR (AIC = −25.50, BIC = −6.54) performed better than models with eGFR (AIC = −18.83, BIC = 2.50). However, the model with eGFR (AIC = 432.50, BIC = 453.83) showed improved fit versus the model without eGFR (AIC = 435.52, BIC = 454.48) for pTau231, supporting its inclusion as a predictor for this analyte despite the smaller sample. Based on these model testing results, we utilized the full sample with longitudinal data to examine pTau217 and pTau181 as outcomes (N=272), and the smaller sample for pTau231 (N=70). Results were considered significant at unadjusted p-value<.05.

## 3. RESULTS

Participants in this study were mostly female (67.6%), White (95.8%), and cognitively unimpaired (97.2%). Of these participants, 50 (17.4%) individuals were amyloid PET positive at the time of the first plasma sample, and 37.6% were carriers of at least one apolipoprotein E epsilon 4 allele (*APOE* ε4). Moreover, 33.1% met obesity criteria based on BMI ≥30 kg/m² and 47.8% were classified as high WHR (men: >0.90, women: >0.85). Among individuals classified as having high BMI, 59.7% were also classified as having high WHR. Cohen’s κ indicates a slight, though significant, overlap between BMI and WHR classifications (κ = 0.14, p<.05).

Twelve individuals (4.2%) had T2DM at baseline, while 6 developed T2DM in subsequent visits (see **Table 1** for participant characteristics). Mann-Whitney U results indicated that plasma pTau analyte concentrations did not differ based on diabetes status or obesity status measured by binarized BMI or WHR at baseline. Partial Spearman’s rho analysis showed that all pTau analytes were significantly correlated, exhibiting large effect sizes (magnitude of the correlations^24^). Plasma pTau181 had the largest effect size with both pTau231 and pTau217 (both relationships Spearman ρ=.63) followed by plasma pTau217 and pTau231 (ρ=.54).

**Table 1.**
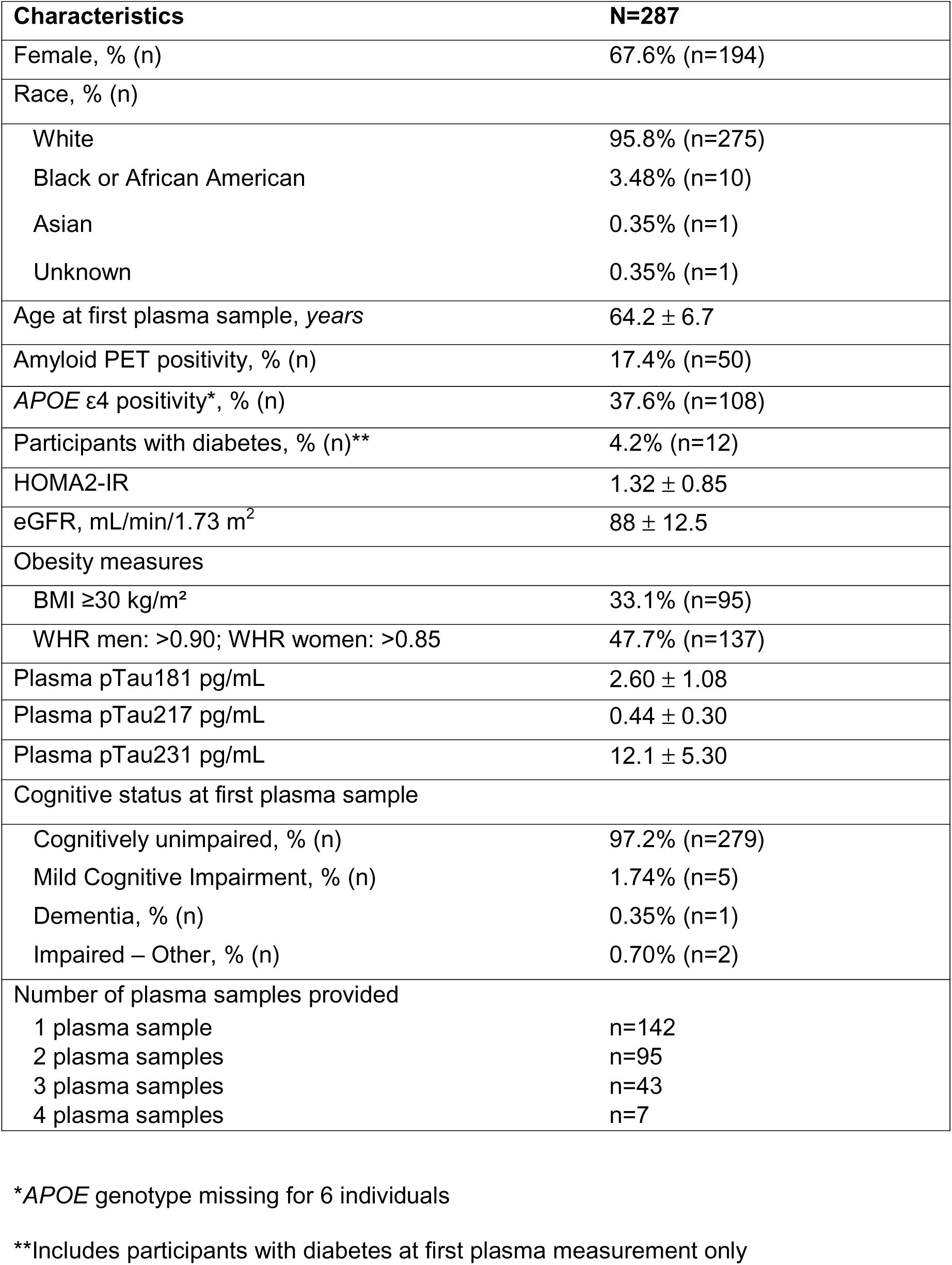
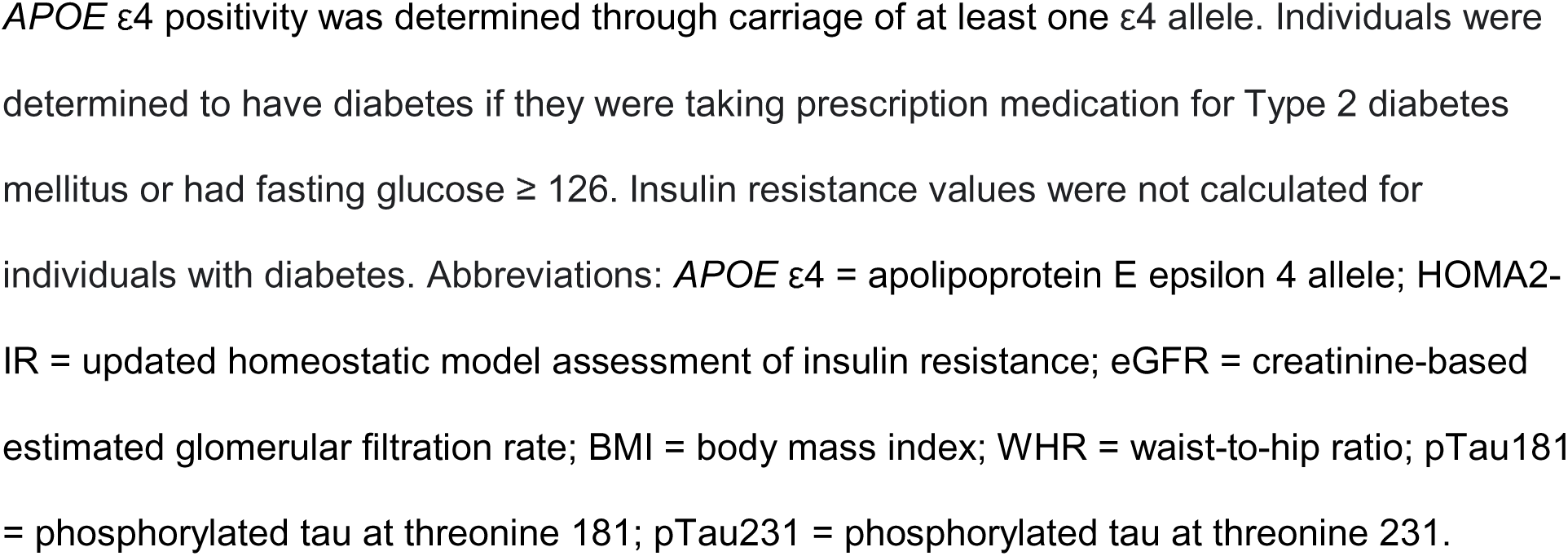
Participant characteristics. Values are provided as mean±standard deviation unless noted otherwise and represent characteristics at the time of the first plasma measurement only.

HOMA2-IR and pTau181 showed a small but significant effect size (ρ=.13, p<.05). Finally, eGFR had a small but significant correlation with plasma pTau217 (ρ=.14, p<.05; **Figure 1**).

**Figure 1.**
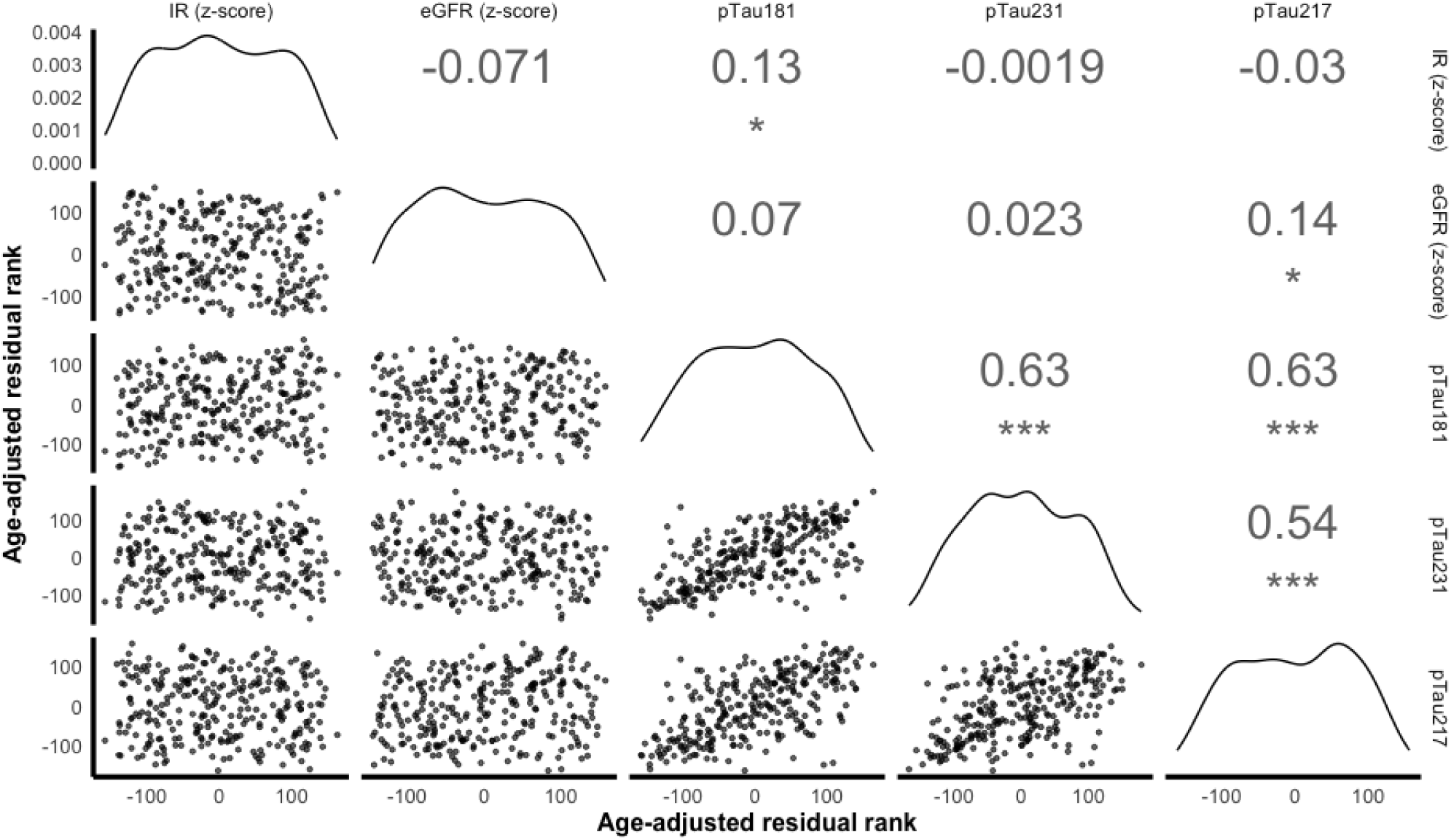
Partial Spearman’s Rho Correlations Matrix. Partial Spearman’s rho correlations were computed to assess the strength and directionality of associations between plasma pTau analytes (pTau181, pTau217, and pTau231) and insulin resistance and kidney function after adjusting for age. Insulin resistance was calculated using the updated homeostatic model assessment of IR (HOMA2-IR) and kidney function was estimated from serum creatinine concentrations using the 2021 CKD-EPI formula to derive estimated glomerular filtration rate (eGFR). Results showed that all plasma pTau analytes were significantly correlated with large effect sizes. Plasma pTau181 was significantly correlated with HOMA2-IR (ρ=.13, p<.05). Plasma pTau217 was correlated with eGFR exhibiting a small effect size (ρ=.14, p<.05). *p<.05 ***p<.001

ROC analysis demonstrated that all plasma pTau analytes were strong classifiers of amyloid positivity. Plasma pTau231 and pTau181 had very similar AUCs (pTau231: AUC=0.78 (95% CI = .071, 0.84); pTau181: AUC = 0.79 (95% CI = 0.74, 0.85)), whereas pTau217 had a larger AUC of 0.91 (95% CI = 0.86, 0.97). Our results indicated that accounting for IR did not improve AUCs for any of the pTau analytes. Finally, accounting for eGFR improved accuracy for both pTau181 and pTau231, but not pTau217 (**Figure 2**).

**Figure 2.**
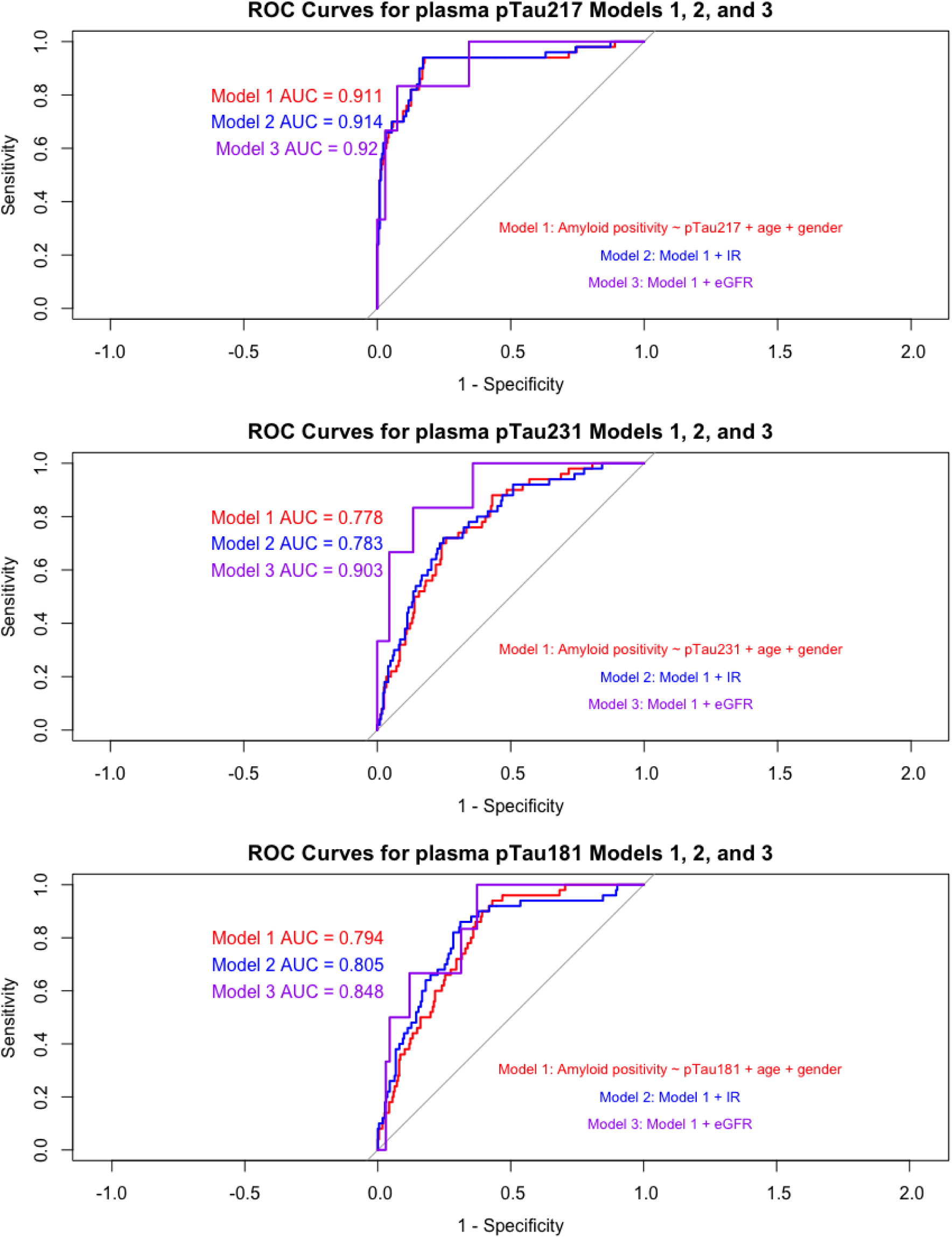
ROC analysis of plasma pTau analytes as predictors of amyloid PET positivity. Receiver operating characteristic (ROC) curves are shown for logistic regression models assessing the concordance of plasma pTau217 (top panel), pTau231 (middle panel), and pTau181 (bottom panel) with amyloid PET positivity. For all models, Model 1 included the plasma pTau analyte, mean-centered age, and gender, Model 2 included Model 1 predictors and HOMA2-IR z-scored, and Model 3 included Model 1 with estimated glomerular filtration rate (eGFR) z-scores. The area under the curve (AUC) was calculated for each model and 95% confidence intervals were estimated. All analysis were completed in R version 4.4.2. Results demonstrated that all plasma pTau analytes were concordant with amyloid positivity with pTau217 showing the highest Model 1 AUC of 0.91 (95% CI = 0.86, 0.97). Accounting for eGFR incrementally improved this accuracy across all pTau biomarkers, however, accounting for IR only incrementally improved accuracy for pTau231 and pTau181, but not pTau217.

Linear mixed models showed that older age significantly predicted mean level of longitudinal pTau181 (β =.04, p<.001), pTau217 (β=0.01, p<0.001), and pTau231 (β=0.20, p<.05). Amyloid positivity status was also associated with mean level of longitudinal pTau181 (β=0.66, p<0.001), pTau217 (β=0.40, p<0.001), and pTau231 (β=4.39, p<.05). For all pTau analytes, high WHR was a significant negative predictor (see **Table 2**). When WHR was modeled continuously, the relationship with pTau181 remained significant (β=-0.15, p<.05), the effect with pTau217 was marginal (β=-0.03, p=.08), and there was no longer an association with pTau231. The direction of all relationships remained in the negative direction, but there was variation in magnitude and significance (see **Supplementary Table**). Finally, the relationship between eGFR and mean level of longitudinal pTau231 was significant (β=−1.01, p<.05) suggesting that lower kidney function was associated with higher levels of pTau231. No other predictors reached significance.

**Table 2.** LME Results for pTau Analytes.

| *Predictors* | **pTau181** |  | **pTau217** |  | **pTau231** |  |
| --- | --- | --- | --- | --- | --- | --- |
|  | *Estimates* | *CI* | *Estimates* | *CI* | *Estimates* | *CI* |
| (Intercept) | 2.54 *** | 2.39 – 2.70 | 0.40 *** | 0.35 – 0.44 | 11.61 *** | 10.28 – 12.95 |
| IR | 0.09 | -0.00 – 0.19 | -0.01 | -0.04 – 0.01 | 0.40 | -0.45 – 1.25 |
| WHR | -0.32 ** | -0.53 – -0.12 | -0.06 * | -0.12 – -0.00 | -1.77 * | -3.51 – -0.03 |
| Age | 0.04 *** | 0.02 – 0.05 | 0.01 *** | 0.01 – 0.01 | 0.20 ** | 0.05 – 0.34 |
| Gender | 0.24 | -0.02 – 0.49 | 0.04 | -0.03 – 0.11 | -0.11 | -2.24 – 2.01 |
| Amyloid status | 0.66 *** | 0.42 – 0.89 | 0.40 *** | 0.33 – 0.46 | 4.39 ** | 1.43 – 7.36 |
| eGFR |  |  |  |  | -1.01 * | -1.88 – -0.13 |
| **Random Effects** |  |  |  |  |  |  |
| $\sigma^2$ | 0.40 | | 0.04 | | 3.20 | |
| $\tau_{00}$ | 0.57 Subject | | 0.04 Subject | | 10.76 Subject | |
| ICC | 0.59 |  | 0.54 |  | 0.77 |  |
| N | 272 Subject |  | 272 Subject |  | 70 Subject |  |
| Observations | 463 |  | 463 |  | 79 |  |
| Marginal R<sup>2</sup> /<br>Conditional R<sup>2</sup> | 0.164 / 0.654 |  | 0.321 / 0.689 |  | 0.320 / 0.844 |  |
| AIC | 1242.203 |  | 102.476 |  | 432.504 |  |
| log-Likelihood | -613.101 |  | -43.238 |  | -207.252 |  |

\* $p < 0.05$ \*\* $p < 0.01$ \*\*\* $p < 0.001$
Three linear mixed models were estimated for each pTau analyte. For pTau181 and pTau217, the models included random intercepts with fixed effects of insulin resistance (IR) z-scored, waist-to-hip-ratio (WHR) binarized, mean-centered age, sex, and amyloid status (positive vs. negative). For pTau231, the model was the same with estimated glomerular filtration rate (eGFR) added as a fixed effect. Model comparison (AIC/BIC) indicated that models without eGFR performed better for pTau181 and pTau217 whereas the model with eGFR performed better for pTau231. All models were fitted in R version 4.4.2 using the lme4 package. Results indicate that WHR, age, and amyloid status were all significant predictors of longitudinal pTau
concentrations for all analytes. For pTau231, eGFR was a significant predictor, indicating that lower kidney function was associated with higher plasma pTau231 concentrations over time.

## 4. DISCUSSION

This study examined the association between metabolic factors (IR, T2DM, BMI, WHR, and eGFR) and plasma pTau biomarkers (pTau181, pTau217, and pTau231) in a head-to-head sense, allowing for a controlled evaluation of relative diagnostic performance under consistent biological conditions. ROC analyses indicated that accounting for IR did not alter concordance of any plasma pTau analyte with amyloid PET, and accounting for eGFR improved concordance with amyloid PET for pTau231 and pTau181 but not pTau217. We observed that eGFR had a small but significant correlation with pTau217 after adjusting for age. Inclusion of eGFR in the ROC analysis, however, did not affect plasma pTau217 concordance with amyloid PET. These findings suggest that pTau217 may be impacted by kidney function in individuals with normal to mildly decreased kidney, but not enough to override its usability as a biomarker for AD-related amyloid pathology. These findings largely align with prior studies indicating that pTau217 may be a more sensitive peripheral marker of AD-related amyloid pathology, particularly in preclinical and prodromal phases^6,25^. Our longitudinal analysis indicated that age, binarized WHR, and amyloid status were all significant predictors of mean level of all plasma pTau analytes, and that lower eGFR associated with higher plasma pTau231. Together, these results indicate that metabolic factors may contribute to pTau variability as measured in plasma, and that such values may need to be interpreted in the context of metabolic conditions.

Baseline HOMA2-IR correlated with pTau181 after adjusting for age, although accounting for IR did not improve the concordance between pTau181 and amyloid PET. It is unclear in our study if the relationship between IR and pTau181 reflects an important biological relationship relevant to dementia risk, or whether IR is operating as a confound. IR can be broadly defined as the inability of tissues in the body to respond to insulin secretion^26^ and is a key feature of hyperinsulinemia (sustained excess blood insulin levels^27^) and T2DM^28^. Insulin is integral for regulating glucose metabolism in the body, and insulin signaling is critical to several neuronal processes^29^. Moreover, IR has been associated with increased risk for AD dementia^30^ and insulin signaling pathways can influence AD-relevant enzymes, including glycogen synthase kinase 3 beta (like GSK-3β), which phosphorylates tau. Hyperinsulinemia can exacerbate the activity of GSK-3β, leading to increased tau phosphorylation and aggregation. We have previously shown that IR is associated with early brain changes in asymptomatic individuals at increased risk for AD, including brain glucose metabolism ^31,32^, amyloid deposition ^33^, and altered brain myelin content^34^. However, when examined directly in relation to brain pathology as assessed with PET, Ennis et al^35^ did not find an association between IR and tau burden in the entorhinal cortex. Although accounting for IR did not improve AUC for any plasma pTau analyte, considering its correlation with pTau181 when interpreting this analyte may be advisable.

Our finding that high WHR was associated with lower mean level of longitudinal plasma pTau concentrations for all analytes may reflect the broader impact of central adiposity on systemic and metabolic health. When modeled continuously, the direction of relationships remained unchanged, although the magnitude and significance of these relationships varied (see **Supplementary Table**). This may indicate that utilizing established cutoffs for WHR may capture non-linear relationships not evident when WHR is modeled continuously, and overall, these results do suggest an effect of WHR on plasma pTau concentrations. Obese individuals are shown to have greater overall total blood volume (e.g., Cepeda-Lopez et al^36^) and the lower plasma concentrations of pTau may be due simply to this effect, although more investigations are needed to understand this phenomenon. Furthermore, as discussed in Rubino et al^19^, obesity, IR, and renal function bidirectionally interact and impacts multiple organ systems.

These complex and integrated processes may impact peripheral clearance mechanisms of pTau proteins and suggests that body fat could confound or moderate plasma pTau interpretations although more research is needed to bring clarity. For plasma pTau181 specifically, our finding contrasts with those from Duff et al^37^, who examined AD plasma biomarker-related changes in individuals after SARS-CoV-2 infection. They found that WHR was associated with higher plasma pTau181 concentrations irrespective of SARS-CoV-2 status, indicating that central adiposity may contribute independently to elevated plasma tau pathology. One possible explanation for this difference could be that our study was conducted in a middle-aged cohort that is relatively healthy, with only 8 reaching the criteria for cognitive impairment. In addition, we did not consider SARS-CoV-2 infection as a covariate, which is relevant given its association with plasma pTau181 concentrations. Not much is currently known about the associations between WHR and plasma pTau231 and pTau217, underscoring the need for further investigation.

A small subsample of participants had available eGFR data for analysis, so these findings, while informative, should be interpreted with caution and as exploratory. Our analysis with eGFR showed that lower eGFR was associated with higher plasma pTau231 concentrations after adjusting for age, sex, WHR, and amyloid status. Moreover, accounting for eGFR resulted in improved AUCs for both pTau181 and pTau231. These findings suggest that even within the range of mildly decreased to normal kidney function, renal clearance may have an impact on plasma pTau concentrations. This contrasts with Arslan et al^38^, who reported no independent associations between eGFR and plasma pTau181, pTau231, or pTau217 after adjusting for age, sex, and amyloid status in a sample from the TRIAD cohort. Possible explanations for this discrepancy include differences in health status or the age distribution as the TRIAD cohort included older participants^38^. Our results altogether reinforce the importance of considering kidney function, even in the absence of CKD, when using pTau biomarkers as an indicator for amyloid pathology, especially for pTau181 and pTau231. This is consistent with prior studies indicating populations with CKD display higher levels of plasma pTau concentrations, possibly due to impaired renal clearance. For example, in a community-dwelling sample from the Mayo Clinic Study on Aging, Mielke et al^39^ found that plasma pTau181 and pTau217 concentrations were higher in individuals with a history of CKD, myocardial infarction, and stroke when adjusting for age, sex, and amyloid PET. Another recent study from the Mayo Clinic showed that individuals with CKD who were amyloid PET-negative displayed higher concentrations of plasma pTau217^40^. Moreover, a study following a sample of participants with CKD before and after kidney transplantation showed that, prior to transplantation, plasma pTau181 levels were elevated in CKD patients relative to controls. After kidney transplantation, plasma pTau181 levels dropped by approximately 71% at 12 weeks, suggesting that elevated pTau181 in CKD likely reflects renal dysfunction as opposed to AD pathology^12^.

The mechanism for how brain pTau fragments is secreted into the bloodstream, and later cleared in the periphery, is not fully known; murine studies may shed light on this process. In one study, Shi et al^41^ injected ^131^I-radiolabeled human 2N4R tau into tau knock-out mice and tracked the physiological efflux of tau from the brain to the blood using fluorescence nanoparticle tracking and found tau-positive exosomes in tau-injected mice but not saline-injected control mice. However, when they looked at clinical samples from humans, they identified more CNS-derived exosomes with tau in the plasma of individuals with Parkinson’s disease versus individuals with AD^41^. In another study, Wang et al^42^ injected ^131^I-radiolabeled tau into cisterna magna of wild-type mice and showed that one-hour after injection the radiolabeled tau distributed to the blood, liver, and mainly kidneys of the animals. Findings from these studies suggest that tau travels from the brain to blood by way of CNS-derived exosomes, that this transport may be impacted by neurodegenerative disease etiology^41^, and that the kidney and liver may be responsible for peripheral tau clearance^42^.

The process by which tau abnormally hyper-phosphorylates, detaches from microtubules, and is cleared into the peripheral bloodstream involves several processes that are ultimately affected by physiological pathways. As such, conditions that alter peripheral metabolism or organ function such as IR, central adiposity, or kidney function, may affect the level of tau fragments measured in plasma. These factors should be carefully considered especially when interpreting plasma pTau concentrations as a proxy for central AD-related amyloid pathology.

### 4.1 Limitations

A few limitations should be noted. First, the participants in this study were mostly white, highly educated, and relatively healthy, limiting generalizability of our sample to the larger population. Our correlation and ROC analysis are cross-sectional in nature, and causation cannot be inferred from these analyses. In addition, this study utilized non-low-molecular weight assay designs, and low molecular weight tau assays may perform differently (e.g., Janelidze et al, 2025^43^). As such, we cannot determine if these metabolic factors we investigated will impact low molecular weight tau assays in similar ways. Although our longitudinal analysis highlighted important associations between age, WHR, amyloid status, and eGFR with pTau231, this analysis was conducted with a limited subset, which could influence robustness of the results for this analyte. Finally, additional studies are needed to determine how brain-derived tau is cleared in the periphery, given that diseases that impact clearance could affect measurements of brain-derived proteins.

### Funding Statement

This work was supported in part by the National Institute on Aging of the National Institutes of Health (NIH) under award numbers F31AG087651 (KLY), R01AG037639 (BBB), R01AG027161 (SCJ), and P50 AG033514, and the University of Wisconsin Institute for Clinical and Translation Research grant 1UL1RR025011. This content is solely the responsibility of the authors and does not necessarily represent the official views of the NIH.

## Data Availability

All data produced in the present study are available upon reasonable request to the authors.

## Acknowledgements

KB is supported by the Swedish Research Council (#2017-00915 and #2022-00732), the Swedish Alzheimer Foundation (#AF-930351, #AF-939721, #AF-968270, and #AF-994551), Hjärnfonden, Sweden (#ALZ2022-0006, #FO2024-0048-TK-130 and FO2024-0048-HK-24), the Swedish state under the agreement between the Swedish government and the County Councils, the ALF-agreement (#ALFGBG-965240 and #ALFGBG-1006418), the European Union Joint Program for Neurodegenerative Disorders (JPND2019-466-236), the Alzheimer’s Association 2021 Zenith Award (ZEN-21-848495), the Alzheimer’s Association 2022-2025 Grant (SG-23-1038904 QC), La Fondation Recherche Alzheimer (FRA), Paris, France, the Kirsten and Freddy Johansen Foundation, Copenhagen, Denmark, Familjen Rönströms Stiftelse, Stockholm, Sweden, and an anonymous filantropist and donor. HZ is a Wallenberg Scholar and a Distinguished Professor at the Swedish Research Council supported by grants from the Swedish Research Council (#2023-00356, #2022-01018 and #2019-02397), the European Union’s Horizon Europe research and innovation programme under grant agreement No 101053962, Swedish State Support for Clinical Research (#ALFGBG-71320), the Alzheimer Drug Discovery Foundation (ADDF), USA (#201809-2016862), the AD Strategic Fund and the Alzheimer’s Association (#ADSF-21-831376-C, #ADSF-21-831381-C, #ADSF-21-831377-C, and #ADSF-24-1284328-C), the European Partnership on Metrology, co-financed from the European Union’s Horizon Europe Research and Innovation Programme and by the Participating States (NEuroBioStand, #22HLT07), the Bluefield Project, Cure Alzheimer’s Fund, the Olav Thon Foundation, the Erling-Persson Family Foundation, Familjen Rönströms Stiftelse, Familjen Beiglers Stiftelse, Stiftelsen för Gamla Tjänarinnor, Hjärnfonden, Sweden (#FO2022-0270), the European Union’s Horizon 2020 research and innovation programme under the Marie Skłodowska-Curie grant agreement No 860197 (MIRIADE), the European Union Joint Programme – Neurodegenerative Disease Research (JPND2021-00694), the National Institute for Health and Care Research University College London Hospitals Biomedical Research Centre, the UK Dementia Research Institute at UCL (UKDRI-1003), and an anonymous donor. BBB receives funding from the National Institute on Aging (NIA) through multiple grants, including R01AG062285, P30AG062715, R01AG037639, R01AG054059, R01AG070883, RF1AG057784, R01AG059312, and R01AG070973.

## Conflicts of Interest

The following authors report no financial or nonfinancial disclosures: KLY, YM, GE, RERR, and NA. EJ receives salary support from the NIH. SCJ serves as a consultant to Eli Lilly, AlzPath, Merck, and Enigma Biomedical. KB has served as a consultant and at advisory boards for Abbvie, AC Immune, ALZPath, AriBio, Beckman-Coulter, BioArctic, Biogen, Eisai, Lilly, Moleac Pte. Ltd, Neurimmune, Novartis, Ono Pharma, Prothena, Quanterix, Roche Diagnostics, Sunbird Bio, Sanofi and Siemens Healthineers; has served at data monitoring committees for Julius Clinical and Novartis; has given lectures, produced educational materials and participated in educational programs for AC Immune, Biogen, Celdara Medical, Eisai and Roche Diagnostics; and is a co-founder of Brain Biomarker Solutions in Gothenburg AB (BBS), which is a part of the GU Ventures Incubator Program, outside the work presented in this paper. HZ has served at scientific advisory boards and/or as a consultant for Abbvie, Acumen, Alector, Alzinova, ALZpath, Amylyx, Annexon, Apellis, Artery Therapeutics, AZTherapies, Cognito Therapeutics, CogRx, Denali, Eisai, Enigma, LabCorp, Merck Sharp & Dohme, Merry Life, Nervgen, Novo Nordisk, Optoceutics, Passage Bio, Pinteon Therapeutics, Prothena, Quanterix, Red Abbey Labs, reMYND, Roche, Samumed, ScandiBio Therapeutics AB, Siemens Healthineers, Triplet Therapeutics, and Wave, has given lectures sponsored by Alzecure, BioArctic, Biogen, Cellectricon, Fujirebio, LabCorp, Lilly, Novo Nordisk, Oy Medix Biochemica AB, Roche, and WebMD, is a co-founder of Brain Biomarker Solutions in Gothenburg AB (BBS), which is a part of the GU Ventures Incubator Program, and is a shareholder of MicThera (outside submitted work). BBB has received consulting fees from New Amsterdam, Cognito Therapeutics, and Merry Life Biomedical. BBB is the founder of Cognovance. Support for meeting attendance includes funding from the Alzheimer’s Association. BBB has served on advisory boards, including the Weston Advisor Grant, the Rush ADRC External Advisory Board, and the Emory ADRC External Advisory Board. Amyloid and tau PET tracers and precursors have been received previously from AVID Radiopharmaceuticals.

## Notes

### Author Declarations

The Institutional Review Board of the University of Wisconsin-Madison gave ethical approval for this work.

