## Supplemental Table for "Metabolic factors associated with plasma phosphorylated tau analytes used to detect Alzheimer’s disease pathology"

| **Supplementary LME Results for pTau Analytes** | | | | | | |
| --- | --- | --- | --- | --- | --- | --- |
|  | **pTau181** | | **pTau217** | | **pTau231** | |
| *Predictors* | *Estimates* | *CI* | *Estimates* | *CI* | *Estimates* | *CI* |
| (Intercept) | 2.38 ^***^ | 2.23 – 2.53 | 0.36 ^***^ | 0.32 – 0.41 | 10.71 ^***^ | 9.44 – 11.98 |
| IR | 0.09 | -0.01 – 0.19 | -0.01 | -0.04 – 0.01 | 0.37 | -0.51 – 1.24 |
| WHR | -0.15 ^*^ | -0.27 – -0.02 | -0.03 | -0.07 – 0.00 | -0.73 | -1.80 – 0.34 |
| Age | 0.04 ^***^ | 0.02 – 0.05 | 0.01 ^***^ | 0.01 – 0.01 | 0.21 ^**^ | 0.06 – 0.36 |
| Gender | 0.27 | -0.01 – 0.56 | 0.05 | -0.03 – 0.13 | -0.03 | -2.38 – 2.32 |
| Amyloid status | 0.64 ^***^ | 0.41 – 0.87 | 0.39 ^***^ | 0.32 – 0.46 | 4.22 ^**^ | 1.14 – 7.30 |
| eGFR |  |  |  |  | -0.94 ^*^ | -1.83 – -0.05 |
| **Random Effects** | | | | | | |
| σ^2^ | 0.40 | | 0.04 | | 3.88 | |
| τ_00_ | 0.58 _Subject_ | | 0.04 _Subject_ | | 10.24 _Subject_ | |
| ICC | 0.59 | | 0.55 | | 0.73 | |
| N | 272 _Subject_ | | 272 _Subject_ | | 70 _Subject_ | |
| Observations | 463 | | 463 | | 79 | |
| Marginal R^2^ / Conditional R^2^ | 0.156 / 0.654 | | 0.318 / 0.692 | | 0.305 / 0.809 | |
| AIC | 1247.699 | | 104.594 | | 435.637 | |
| log-Likelihood | -615.849 | | -44.297 | | -208.819 | |
| ** p<0.05   ** p<0.01   *** p<0.001* | | | | | | |

We estimated three linear mixed models for each pTau analyte. For pTau181 and pTau217, the models included random intercepts with fixed effects of insulin resistance (IR) z-scored, waist-to-hip-ratio (WHR) z-scored, mean-centered age, sex, and amyloid status (positive vs. negative). For pTau231, the model was the same with estimated glomerular filtration rate (eGFR) added as a fixed effect. Results indicate that age and amyloid status were all significant predictors of longitudinal pTau concentrations for all analytes. WHR z-scored was negatively associated with all pTau analytes, however, this relationship was significant for pTau181 (p<.05), marginal for pTau217 (p=.08), and non-significant for pTau231.
